# Metabolic Basis of Post-Infectious Sequelae After Ebola Virus Disease

**DOI:** 10.64898/2026.01.02.25343095

**Authors:** Anna Sanford, Nell Bond, Samuel Ficenec, Charlotte Osterman, Payton Farkas, Emily Engel, Bronwyn Gunn, Donald S. Grant, Robert Samuels, Kevin Zwezdaryk, John Schieffelin

## Abstract

Ebola virus disease (EVD) survivors often present with clinical sequelae after acute disease resolution, called post-Ebola syndrome (PES). Why some survivors develop these sequelae and others do not is poorly defined. Altered metabolism has been noted in acute EVD but not studied in PES. We identified differential expression of metabolites involved in multiple metabolic pathways in EVD survivors with PES. This included the tricarboxylic acid cycle, amino acid, nucleotide, and short chain fatty acid metabolism. Several of these pathways are associated with immune dysfunction. The identified metabolites have potential use as biomarkers of post-Ebola syndrome.

## Background

Many Ebola virus disease (EVD) survivors develop post-Ebola syndrome (PES) which consists of heterogenous sequelae including musculoskeletal, cardiopulmonary, neurologic, and constitutional (headache, fatigue) symptoms often identifiable long after acute illness^1,2,3^. The mechanisms driving the development of these sequelae in some survivors but not others remain unclear. With improvement in the treatment of Ebola virus (EBOV) infected individuals, it is likely there will be an increase in survivors with PES. Therefore, a better understanding of the etiology of PES is crucial for identification of survivors at risk for PES and for future PES treatments.

Metabolomics can elucidate metabolic changes after viral infection and, therefore, suggest markers of post-viral sequelae for further investigation. Despite its utility, metabolomics has been used sparingly in studies of post-viral sequelae, although there has been work focusing on metabolic response in Long COVID^4^ and HIV sequelae^5^. Ebola virus has been shown to alter fatty acid and amino acid metabolism in cells stimulated with virus-like particles^6^, and EVD fatalities have been associated with a decrease in plasma free amino acids^7^. However, metabolic markers of PES have not been described. This raised the question of whether altered metabolism persists in EVD survivors with PES, as well as implications of identified metabolic changes. We hypothesized that EVD survivors with PES would demonstrate a unique metabolic profile compared to asymptomatic survivors and aimed to identify possible biomarkers of PES for further study.

## Methods

### Study Design and Population

A convenience sample of plasma was collected as part of an ongoing cohort study of EVD survivors and household contacts (HCs) in Sierra Leone as previously described^8^. Survivors with musculoskeletal/gastrointestinal (MSK/GI) or cardiopulmonary (CP) sequalae were included in the PES group. Samples were from three previously identified groups-EVD survivors with PES (MSK/GI or CP sequelae), asymptomatic glycoprotein (GP) IgG positive survivors, and asymptomatic GP IgG negative HCs. The HCs were selected to have a similar demographic background to survivors (age, sex, sample collection date) while being from the same region as EVD survivors. Some HCs were from the same households as survivors included in this study while others were from separate households.

### Clinical Characteristics

Survivors in the MSK/GI and CP sequelae groups were identified from our previous studies^8,9^. Sequelae considered MSK/GI sequelae include physical exam signs: decreased joint range of motion, extremity edema or effusion, joint tenderness to palpation, and abdominal tenderness to palpation. We prioritized including survivors with more frequent MSK/GI sequelae, although only one time point per survivor was analyzed in this study. Survivors who presented with multiple cardiopulmonary signs or symptoms were prioritized for inclusion in the CP group. Sequelae considered to be cardiopulmonary in origin included symptoms of chest pain, heart palpitations, shortness of breath, or a cough^9^. Physical exam findings included as cardiopulmonary sequelae were the absence of S1 or S2 heart sounds, the presence of murmurs, rubs or gallops, an abnormal heart rate, an irregular heart rhythm, lower extremity edema, poor distal pulses, or the presence of wheezes, rales, rhonchi on auscultation or dullness to percussion of the posterior chest^9^. Asymptomatic GP IgG+ survivors were defined as asymptomatic based on principal components analysis clustering as defined in our previous study^8^. All survivors with MSK/GI sequelae were grouped into the MSK or MSK/GI cluster at first visit from our previous study. Most of the survivors with cardiopulmonary sequelae were grouped in the cardiac/GI cluster at first visit but three survivors did not cluster into a sequelae group at first visit and were later noted to have cardiopulmonary sequelae. Demographic characteristics and body mass index (BMI) were collected at first visit. If BMI was not collected at first visit, then data from the nearest follow-up visit was used.

### Sample and Data Collection

Plasma samples were collected from March 2016 to April 2019. Clinical data were collected as close to plasma collection date as possible (Supplementary Table 1). Blood samples were collected by venipuncture and plasma was processed according to standard methods. Using LC-MS/MS, untargeted metabolomics was completed for each sample by Gigantest Inc. (Baltimore, MD). Extraction was performed using cold acetonitrile and formic acid. Metabolites were identified using in-house databases and quantified using XCalibur software for data analysis against authenticated external standards recorded under identical conditions.

### Data Analysis

Univariate, multivariate, biomarker and pathway analyses were performed in MetaboAnalyst 6.0^10^. Peak intensities were used for analysis. Missing values (n=2762, 8.9%) were replaced with 1/5 the minimum positive values of their corresponding variables. Data was filtered using MetaboAnalyst 6.0 presets: variables were removed if they had a relative standard deviation (standard deviation/mean) greater than 25% and 25% of variables were removed based on interquartile range (IQR), selecting for near constant values (lower tail of IQR). Data was normalized to median and log transformed (base 10). Logistic regression determined whether demographic characteristics predicted group membership (PES vs asymptomatic survivor) using R Studio version 4.4.2 (Boston, MA). Significantly different metabolites on univariate analysis were defined as fold change (FC) > 2.0 and adjusted p-value (FDR via Benjamini-Hochberg (BH) method) <.05 using Student’s t-test. Spearman’s R correlation was performed in JMP, Version 18.1.0. (SAS Institute Inc., Cary, NC, 1989-2025). Network analysis of significant correlations of tricarboxylic acid (TCA) cycle metabolites was performed in Cytoscape Version 3.10.3^11^. Principal components analysis (PCA) and partial least squares discriminant analysis (PLS-DA) were performed in MetaboAnalyst. Variable importance of projection (VIP) scores were calculated from the top 15 PLS-DA metabolites in Components 1 and 2. Significant pathways included in pathway analysis were required to have an impact score >0.2 and FDR <.05. For biomarker analysis, PLS-DA and Random Forest (RF) models were trained with a discovery cohort of 39 randomly selected survivors (approximately 2/3 of survivors). One hundred runs of Monte-Carlo cross validation (MCCV) were performed on the discovery models.

The validation cohort consisted of the remaining 18 randomly selected survivors. Each cohort was randomly assigned a proportional number of survivors with PES and asymptomatic survivors. The top 10 most important metabolites contributing to the discovery model were tested on the validation cohort. Receiver operating characteristic (ROC) curves were generated from 100 cross validated runs for both discovery and validation cohorts. Data visualization was performed in MetaboAnalyst and JMP.

### IRB Determination and Written Consent

This study was approved by the Tulane University Institutional Review Board (IRB approval number: 701226) and the Sierra Leone Ethics and Scientific Review Committee (021/11/2024). Written informed consent was provided by adult participants ≥18 years, consent by a parent or guardian and assent in children 12-17 years, and consent by a parent or guardian in children <12 years. Study personnel were all trained in ethics and research compliance.

## Results

### EVD Survivors with PES Display a Unique Metabolome

EVD survivors with PES (n=37) demonstrated an altered metabolic profile compared to asymptomatic EVD survivors (n=20), characterized largely by downregulation of multiple metabolic pathways (Figure 1A; adjusted p-value for all comparisons: <.05). Age, sex, BMI, and sample collection date were not significant for predicting PES when comparing EVD survivors with PES and asymptomatic survivors (Supplementary Table 2). There were no significantly different metabolites when comparing survivors with MSK/GI sequelae (n=18) to survivors with cardiopulmonary sequelae (n=19). There were no significantly different metabolites when comparing survivors with PES and asymptomatic HCs (n=20).

**Figure 1:**
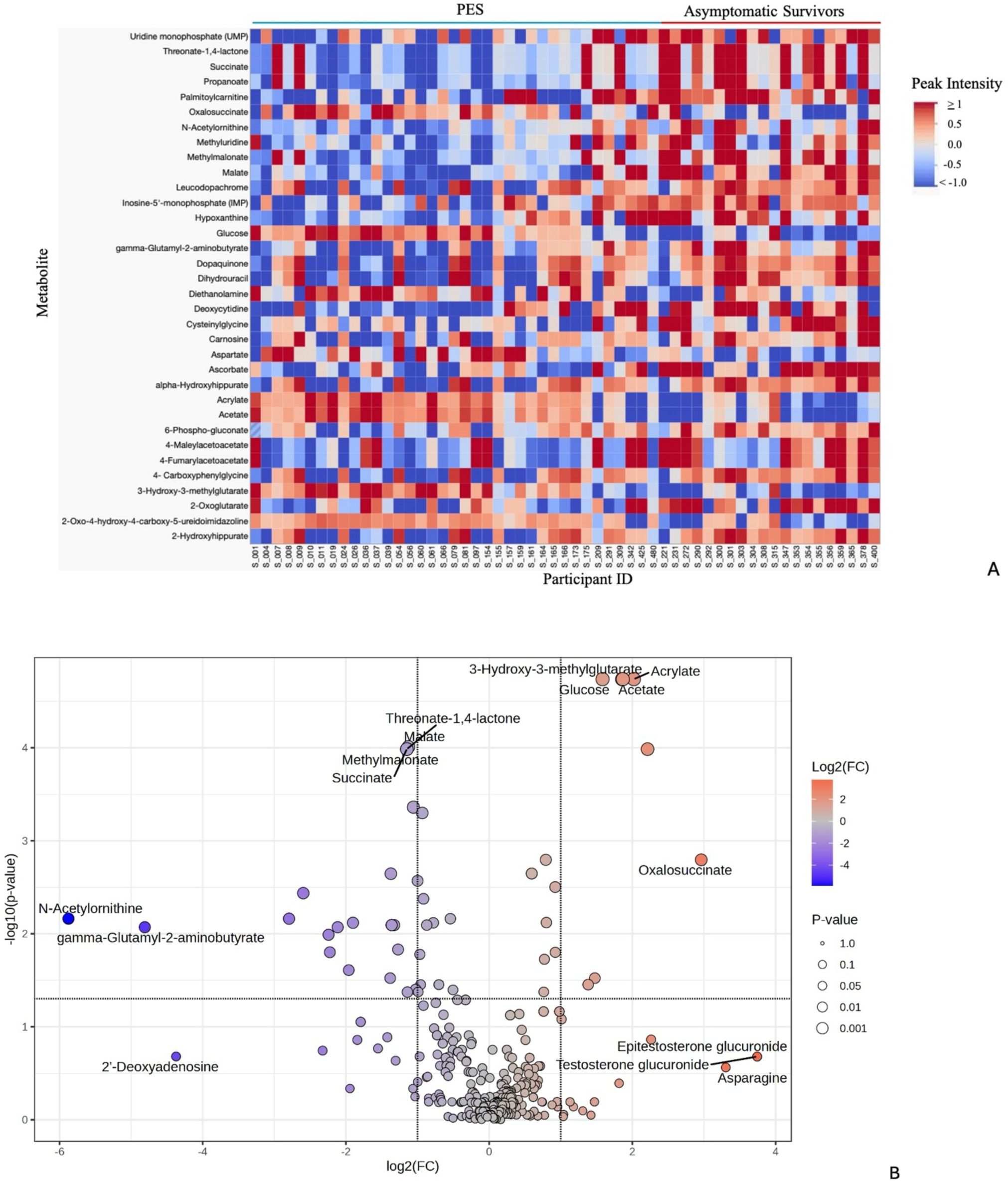
EVD Survivors with PES Demonstrate a Unique Metabolic Profile Compared to Asymptomatic Survivors Figure 1A: 26 metabolites are significantly downregulated in the PES group and 8 metabolites are significantly upregulated compared to asymptomatic survivors. Differential metabolites are defined as FC > 2.0 and adjusted p-value (FDR) <.05. All peak intensities greater than or equal to 1 are illustrated in dark red, all peak intensities equal to or less than −1 are illustrated in dark blue. Analysis was performed in MetaboAnalyst and visualized in JMP. Peak intensities were normalized to median and log transformed. Figure 1B: Volcano plot demonstrates differentially expressed metabolites in EVD survivors with PES compared to asymptomatic survivors. P-value displayed is adjusted p-value (FDR). Analysis and visualization performed in MetaboAnalyst. FC= fold change

Using univariate analysis, five hundred and forty-four metabolites were identified and 34 were differentially expressed when comparing EVD survivors with PES and asymptomatic survivors. Twenty-six metabolites were significantly decreased and eight metabolites were significantly increased in this comparison (Figure 1A-B; Supplementary Table 3). Key pathways demonstrating altered metabolism in EVD survivors with PES compared to asymptomatic survivors included nucleotide metabolism, the TCA cycle, and short chain fatty acid (SCFA) metabolism.

Spearman’s R correlation analysis of EVD survivors with PES demonstrated significant relationships of metabolites in several metabolic pathways (Figure 2A). There was no significant correlation between demographic characteristics (BMI, age, sex) or any metabolite in the PES group; however, BMI significantly correlated with age (positive correlation). Multiple significant correlations of TCA cycle metabolites and metabolites of amino acid metabolism, fatty acid metabolism, and nucleotide metabolism were noted in the PES group (Supplementary Table 4). Given the number of TCA metabolites affected, we further illustrated this pathway via network analysis (Figure 2B).

**Figure 2:**
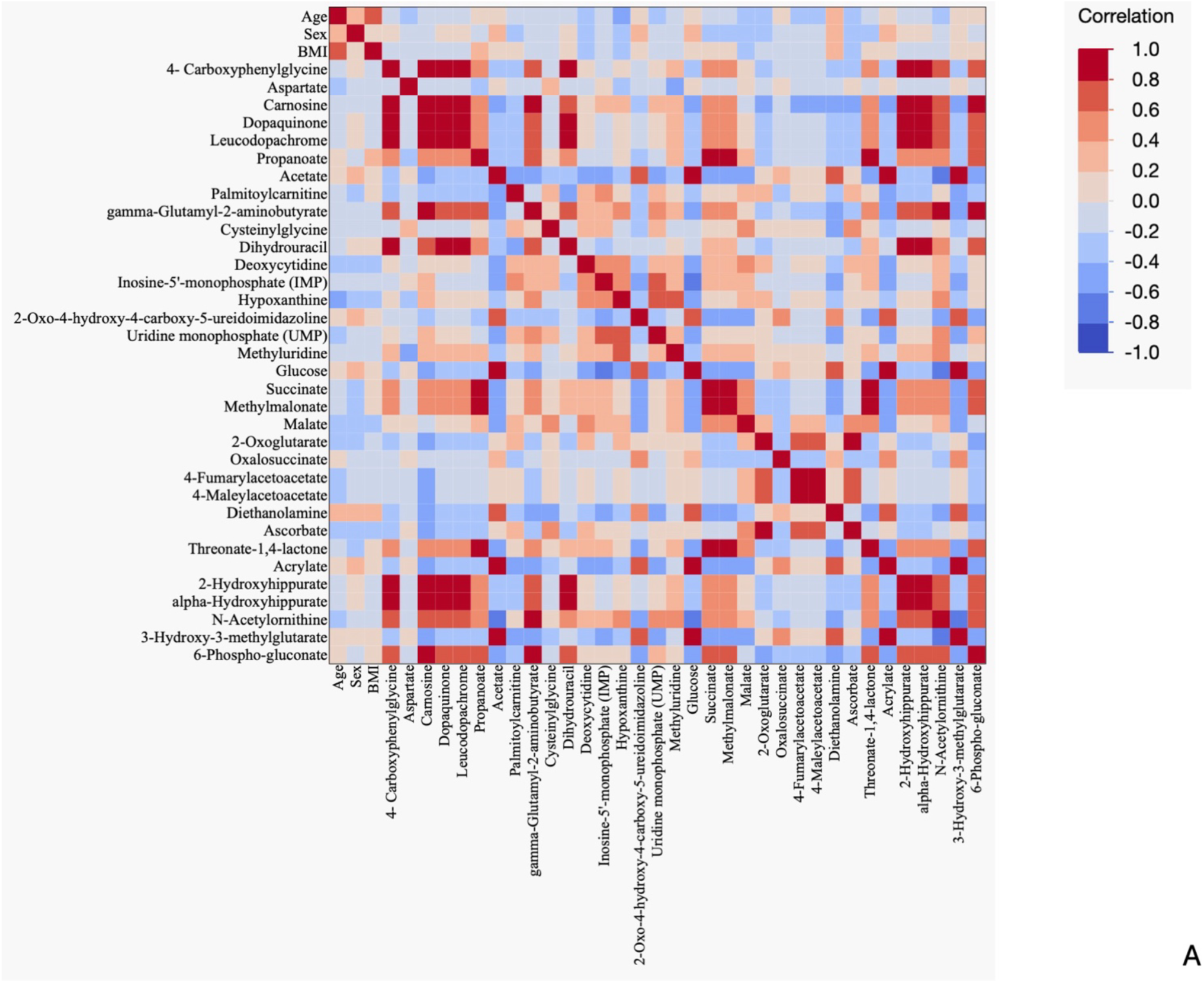
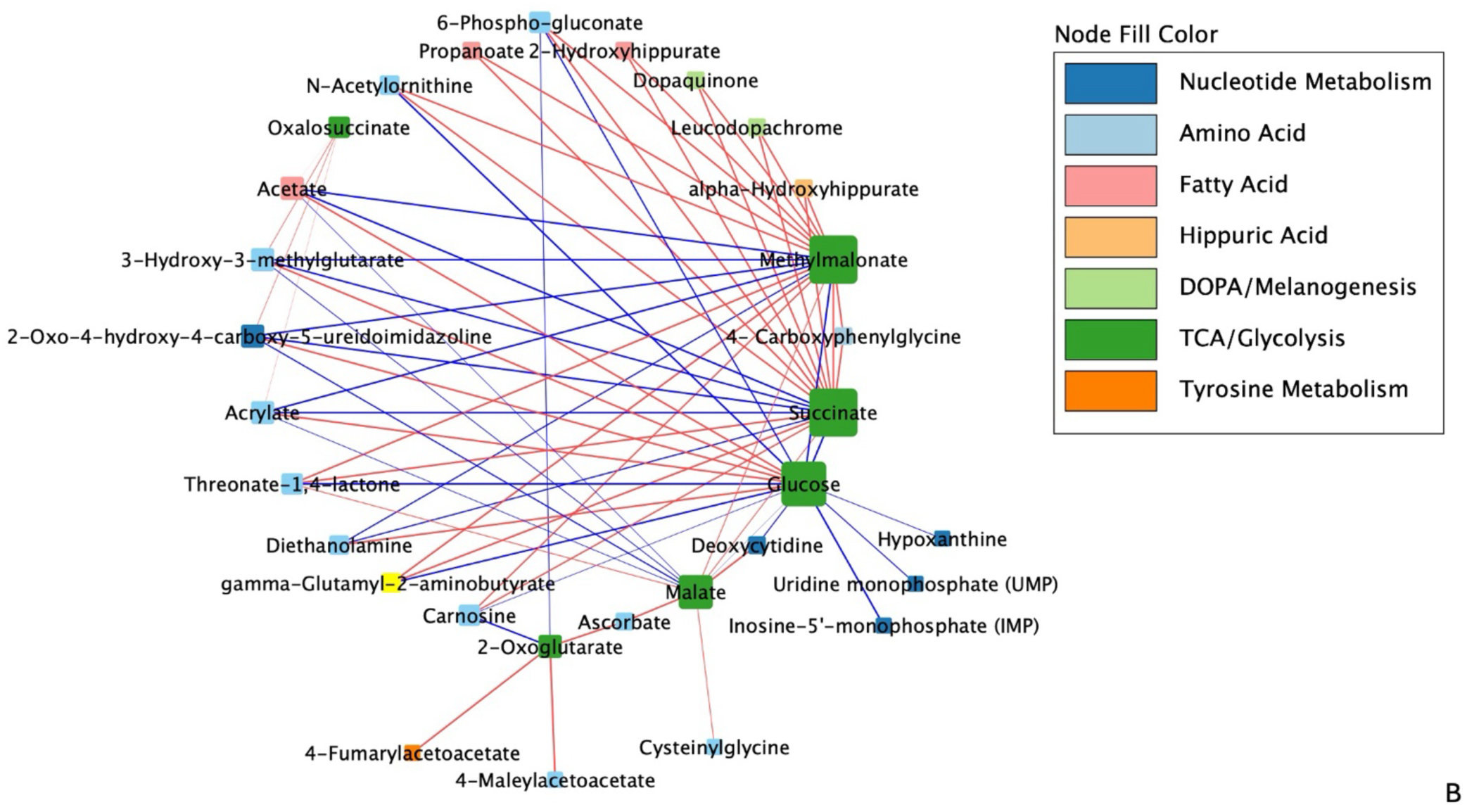
Correlation Analysis Reveals Significant Interactions Between Metabolic Pathways in EVD Survivors with PES Figure 2A: Heatmap of Spearman’s rank correlation analysis of EVD survivors with PES including demographic characteristics (age, sex, BMI) and significantly differential metabolites. Included metabolites were considered significant if FC was >2.0 and FDR <.05 when comparing EVD survivors with PES to asymptomatic survivors. Correlations between variables of survivors with PES are red if positive and blue if negative. Figure 2B: Network analysis of TCA cycle metabolites in EVD survivors with PES. Node size was determined by the number of connections for the metabolite (larger node corresponds to more connections). Node fill color demonstrates which pathway the metabolite belongs to. Line thickness was determined by the p-value of the correlation (smaller p-value corresponds to thicker line). Line color demonstrates whether association is positive (red) or negative (blue). Relationships were determined by Spearman’s rank correlation performed in JMP. Only significant relationships (FDR <.05) were included. Correlation network visualized using Cytoscape. Supplementary table 4 contains rho values.

PCA and PLS-DA showed separation of EVD survivors with PES and asymptomatic survivors based on metabolic profile (Supplementary Figure 1, Figure 3). Metabolites associated with the TCA cycle, SCFA metabolism, and glucose all demonstrated importance for Components 1 and 2 of the PLS-DA model (Supplementary Table 5).

**Figure 3:**
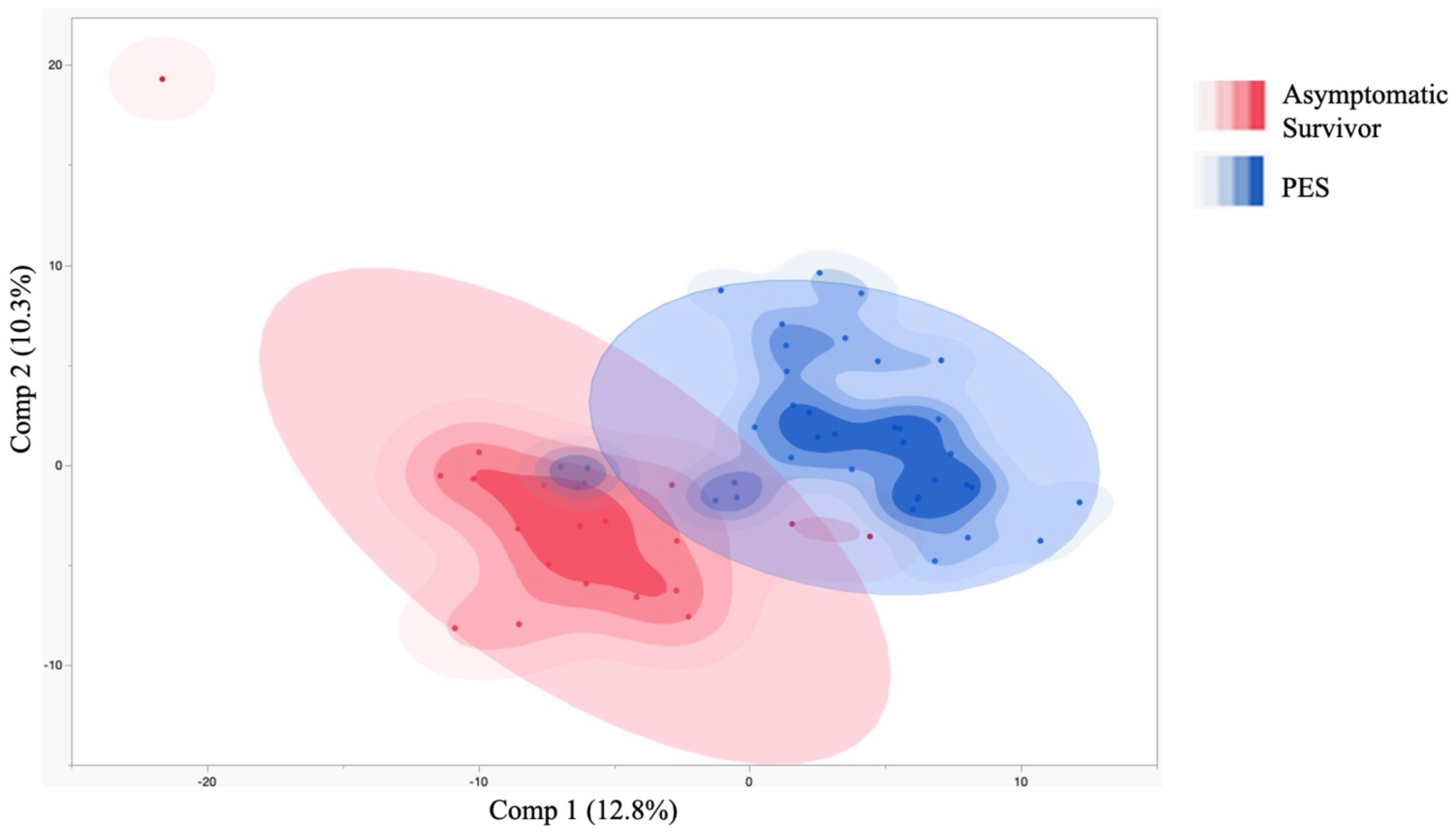
EVD Survivors with PES and Asymptomatic Survivors Separate into Distinct Groups Based on Metabolic Profile Partial least squares discriminant analysis (PLS-DA) demonstrates separation of PES and asymptomatic survivor groups. The highest accuracy PLS-DA model included 2 components with an accuracy of 0.86, R2=0.66, and Q2=0.45. Data analyzed in MetaboAnalyst and visualized in JMP.

### Survivors with PES Demonstrate Downregulation of Multiple Metabolic Pathways

Pathway analysis demonstrated significantly downregulated metabolites in EVD survivors with PES compared to asymptomatic survivors in multiple pathways (Figure 4, FDR <.05). The most impactful pathways involved amino acid metabolism (taurine, hypotaurine, tyrosine, cysteine and methionine, among others). Nucleotide, TCA cycle, glycolysis, and pyruvate metabolism pathways were also significantly downregulated. Fewer pathways were significantly upregulated or were significantly involved with an equal number of up and downregulated metabolites (Supplementary Figure 2). Significantly upregulated pathways included starch and sucrose metabolism, tryptophan metabolism, arginine and proline metabolism, nicotinate and nicotinamide metabolism, pentose and glucuronate interconversions. Individual metabolites from pathway analysis of the TCA cycle are demonstrated in Supplementary Figure 3.

**Figure 4:**
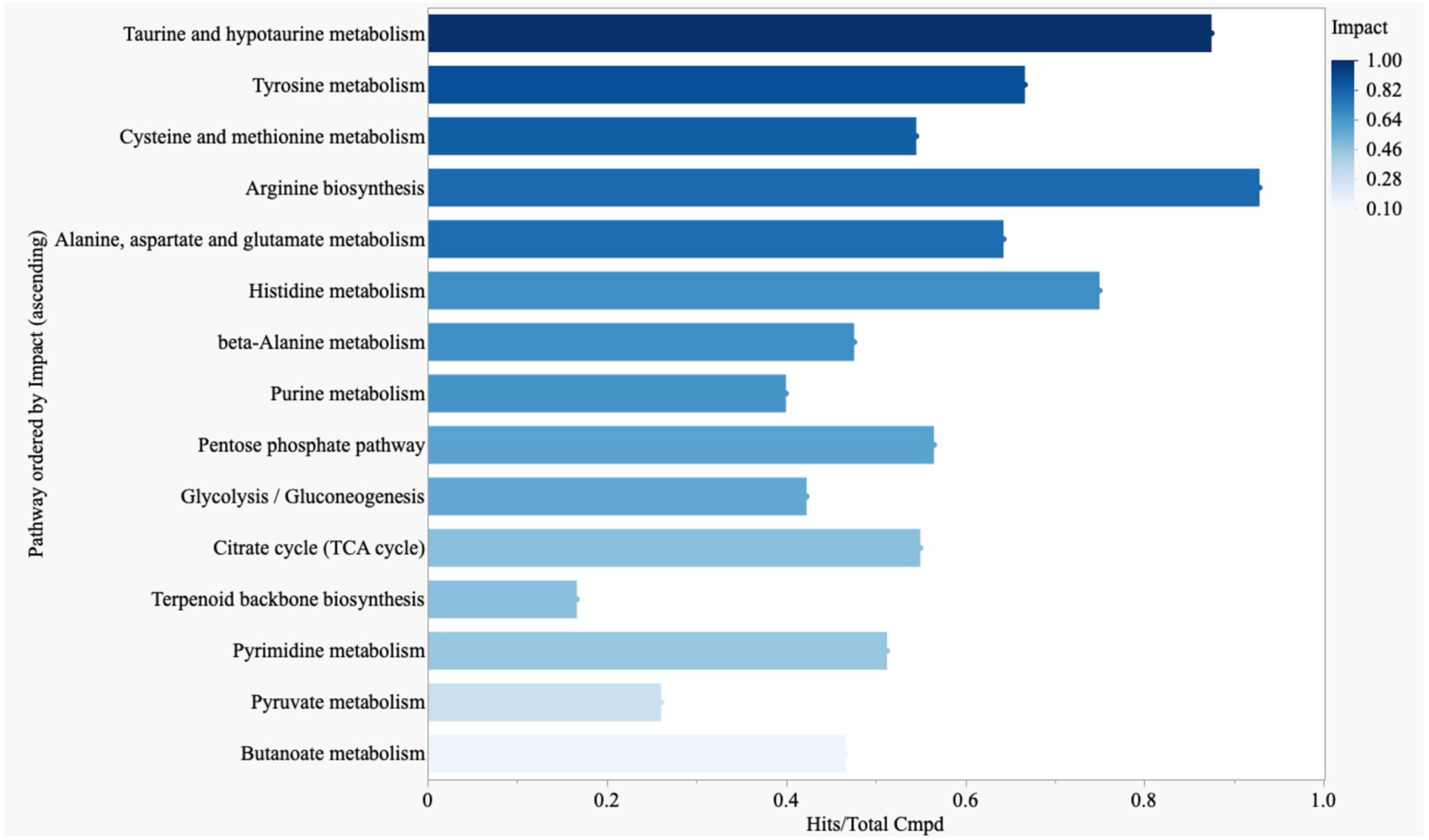
Metabolites of TCA Cycle, Amino Acid, Nucleotide, and SCFA Pathways Characterize the Metabolic Profile in PES Pathway analysis demonstrates significantly impactful pathways in EVD survivors with PES compared to asymptomatic survivors. The included pathways demonstrated primarily downregulated metabolism, determined by most significant metabolites in that pathway being lower in EVD survivors with PES compared to asymptomatic survivors. Pathways ordered by impact score. Impact score required to be >0.2 and FDR <.05 for inclusion. Pathway analysis performed in MetaboAnalyst and data visualized using JMP.

### Biomarker Analysis: Metabolites Predict Post-Ebola Syndrome

Biomarker analysis identified ten metabolites predictive of PES when comparing EVD survivors with PES to asymptomatic survivors. The ten metabolites predictive of PES were the same in the RF and PLS-DA models, both with areas under the curve (AUC) >0.8 (Supplementary Figure 4A-B). These metabolites included SCFA and TCA cycle metabolites, among others. These ten metabolites were also significant in the univariate analysis comparing EVD survivors with PES and asymptomatic survivors (Figure 1A). These predictive metabolites were then used in a separate validation cohort to predict PES and again demonstrated areas under the curve >0.8 (Figure 5A-B). Predictive accuracy for the validation cohort was 0.799 and 0.796 for the for the PLS-DA and RF models, respectively.

**Figure 5:**
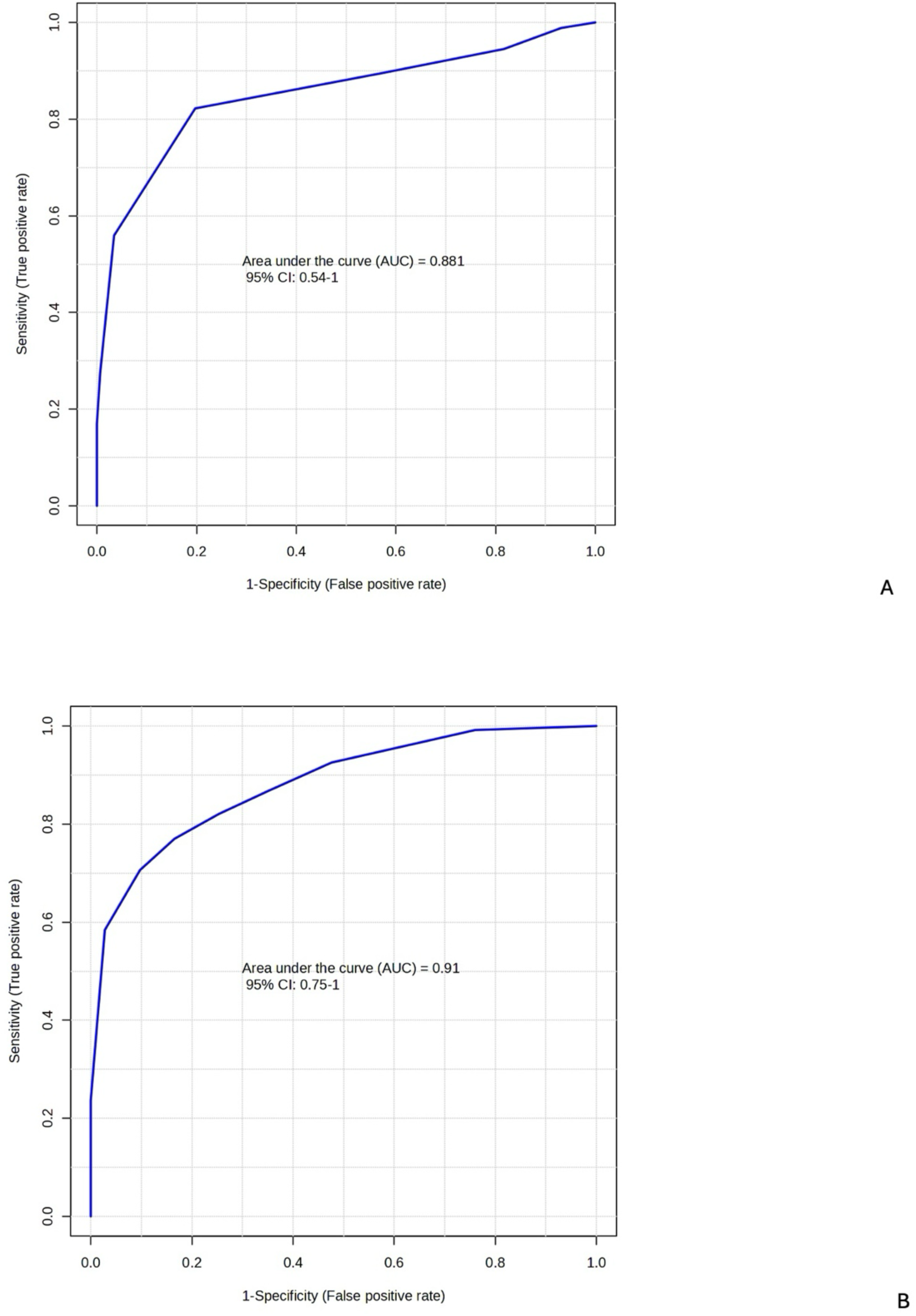
Receiver Operating Characteristics (ROC) Curves Demonstrate Ability of PLS-DA and RF Models to Distinguish Between Ebola survivors with PES and Asymptomatic Survivors. The top ten most important metabolites in both models were acrylate, glucose, acetate, 3-hydroxy-3-methylglutarate, malate, 2-Oxo-4-hydroxy-4-carboxy-5-ureidoimidazoline, succinate, methylmalonate, threonate-1,-4-lactone and propanoate. Data analyzed and visualized in MetaboAnalyst. Discovery models available as Supplementary Figure 4A-B. Figure 5A: PLS-DA validation model. Figure 5B: Random Forests (RF) validation model.

## Discussion

Here, we demonstrate that Ebola survivors with PES, compared to asymptomatic survivors, exhibit downregulated metabolism of multiple metabolic pathways. Downregulation was observed in the TCA cycle, amino acid and nucleotide metabolism pathways. SCFA (propanoate, acetate) were also significantly altered. Metabolites involved in these pathways were identified as potential biomarkers using two additional models: RF and PLS-DA. The same ten metabolites were identified via both analyses, reinforcing the possibility that these metabolites play a role in PES.

Our findings are consistent with metabolic changes seen in cells stimulated with EBOV-like particles^6^ as well as chronic viral infections^12,13^. As previously discussed, amino acid metabolism is affected in cells stimulated with EBOV-like particles^6^. In people living with human immunodeficiency virus (HIV), changes to amino acid metabolism and signs of mitochondrial disruption including TCA cycle dysfunction have been noted in transient controllers of HIV compared to persistent controllers^12^. Altered mitochondrial function has been associated with T-cell exhaustion in chronic infections such as HIV, hepatitis B virus (HBV), and lymphocytic choriomeningitis virus (LCMV)^13^. Our findings could indicate similar immune changes in EVD survivors with PES. While our results are from plasma samples and cannot be directly linked to the cellular level, these findings warrant further investigation.

The changes seen in SCFA among survivors with PES (increased acetate, decreased propanoate) could be due to dysregulation of the gut microbiome in EVD survivors with PES. Reduced SCFA-producing symbionts have been noted in persons with Long COVID^14^. SCFAs can influence immune response, typically encouraging an anti-inflammatory response. SCFAs have also been shown to modulate CD8+ T-cell activity^15^.

Although several studies have focused on immune biomarkers of PES^16,17^, no clear profile has been identified. Prior research in CD8+ T cell activity in PES has demonstrated that more survivors with PES demonstrate an EBOV-specific CD8+ T cell response compared to asymptomatic survivors^16^. As immune function and differentiation are mediated by metabolic pathways, we questioned if Ebola virus disrupts metabolic homeostasis. We observed changes in the TCA cycle that are associated with decreased CD8+ T-cell activity in chronic infections or post-viral syndromes^13^. Additionally, SCFA changes have been associated with other post-viral sequelae^14^ and influence CD8+ T-cell activity^15^.

There are several limitations to our study. First, the sample size is a limitation. It is possible that additional key metabolites associated with PES would be identified in a larger study. Furthermore, there was no significant difference in univariate analysis when comparing EVD survivors with PES and household contacts. This could be due to several reasons, including limited sample size, as trends for several metabolites were similar in this comparison but not statistically significant (data not shown). Another possibility is that EVD survivors without PES demonstrate unique metabolic changes which contribute to the significant difference between EVD survivors with and without PES. As we only studied plasma samples, we can only suggest potential cellular mechanisms of metabolic changes in EVD survivors with PES. Further studies at the cellular level, particularly looking at T-cell function and immunometabolism, are required to determine if the metabolic changes we observed are related to immune dysfunction.

In conclusion, EVD survivors demonstrate altered metabolism in the TCA cycle, amino acid, nucleotide, and SCFA pathways. Several of these pathways have been linked to immune dysfunction (particularly in CD8+ T cells) and chronic infection in studies of other viral infections. Further studies of cellular immunometabolism in EVD survivors with PES are needed. While our work focuses on metabolic changes in a specific group, EVD survivors with PES, we hope our work also demonstrates the utility of metabolomic study of post-viral sequelae more broadly.

## Acknowledgements

This work is dedicated to those affected by Ebola virus disease.

## Author contributions

Conceptualization: AS, NB, KZ, JS; Data curation: NB, EE; Formal analysis: AS, NB, KZ, JS; Funding acquisition: AS, JS; Investigation: CO, PF, RS, AS; Methodology: AS, NB, KZ, BG, JS; Project administration: AS, NB, EE, DG, JS; Resources: NB, SF, EE, RS, BG, DG, JS; Supervision: NB, KZ, JS; Visualization: AS, NB, KZ, JS; Writing-original draft: AS, NB, JS; Writing-review and editing: AS, NB, JS, RS, KZ, SF.

## Data Availability Statement

Raw data is available upon reasonable request to the authors and will be uploaded to an online data repository.

