## Supplementary Tables and Figures for "Metabolic Basis of Post-Infectious Sequelae After Ebola Virus Disease"

**Supplementary Data**

**Supplementary Tables**

| Sample Collection Date (MM-YYYY) | Clinical Sequelae Date (MM-YYYY) | Sequelae |
|---|---|---|
| 03-2016 | 03-2016 | MSK/GI |
| 02-2017 | 03-2016 | MSK/GI |
| 03-2016 | 03-2016 | MSK/GI |
| 03-2016 | 03-2016 | MSK/GI |
| 03-2016 | 03-2016 | CP |
| 02-2017 | 03-2016 | MSK/GI |
| 08-2018 | 03-2022 | CP |
| 04-2019 | 04-2019 | CP |
| 03-2016 | 03-2016 | MSK/GI |
| 02-2017 | 02-2017 | MSK/GI |
| 04-2019 | 04-2019 | CP |
| 04-2019 | 03-2020 | CP |
| 08-2018 | 03-2022 | CP |
| 03-2016 | 03-2016 | MSK/GI |
| 03-2017 | 03-2017 | MSK/GI |
| 03-2017 | 03-2017 | CP |
| 04-2019 | 04-2019 | CP |
| 03-2017 | 03-2017 | MSK/GI |
| 09-2016 | 09-2016 | MSK/GI |
| 03-2016 | 03-2016 | MSK/GI |
| 04-2019 | 03-2020 | CP |
| 04-2019 | 03-2020 | CP |

| | | |
|---|---|---|
| 10-2016 | 04-2017 | CP |
| 04-2017 | 04-2017 | CP |
| 04-2017 | 04-2017 | CP |
| 04-2017 | 04-2017 | CP |
| 04-2016 | 04-2016 | CP |
| 04-2016 | 04-2017 | CP |
| 04-2016 | 04-2016 | MSK/GI |
| 03-2016 | 08-2016 | CP |
| 08-2016 | 08-2016 | CP |
| 07-2017 | 07-2017 | MSK/GI |
| 11-2017 | 11-2017 | MSK/GI |
| 11-2017 | 11-2017 | MSK/GI |
| 12-2017 | 11-2017 | MSK/GI |
| 12-2017 | 11-2017 | MSK/GI |
| 09-2018 | 03-2020 | CP |

Supplementary Table 1: Dates of plasma collection and sequelae for all survivors with sequelae. ID not shown to protect patient privacy.

| | **PES (n=37)** | **Asymptomatic Survivors (n=20)** | **p-value** | **PES & Asymptomatic Survivor (n=57)** |
|---|---|---|---|---|
| **Median age (y)** | 30 | 28 | 0.22 | 29 |
| **Female (%)** | 59.5 | 60 | 0.99 | 59.6 |
| **Median BMI ($kg/m^2$)** | 24.5 | 22.1 | 0.11 | 23.1 |
| **Median Sample Collection Date** | 2017-03-08 | 2017-11-26 | 0.11 | 2017-11-21 |

| | | | | |
|---|---|---|---|---|
| | **PES (n=37)** | **Household Controls (n=20)** | **p-value** | **PES & Household Controls (n=57)** |
| **Median age (y)** | 30 | 29.5 | 0.75 | 30 |
| **Female (%)** | 59.5 | 60.0 | 0.98 | 59.6 |
| **Median BMI ($kg/m^2$)** | 24.5 | 23.5 | 0.49 | 24.4 |
| **Median Sample Collection Date** | 2017-03-08 | 2017-07-14 | 0.50 | 2017-04-24 |
| | **MSK/GI (n=18)** | **CP (n=19)** | **p-value** | **MSK/GI & CP (n=37)** |
| **Median age (y)** | 30 | 25 | 0.82 | 30 |
| **Female (%)** | 66.7 | 52.6 | 0.89 | 59.5 |
| **Median BMI ($kg/m^2$)** | 27.2 | 22.7 | 0.11 | 24.5 |
| **Median Sample Collection Date** | 2017-02-12 | 2017-04-24 | 0.02* | 2017-03-08 |

Supplementary Table 2: Logistic regression included age, sex, BMI and sample collection date to predict group membership. No characteristic was significant in predicting PES when comparing EVD survivors with PES and asymptomatic survivors or EVD survivors with PES and household controls. EVD survivors with cardiopulmonary (CP) sequelae had a significantly later collection date compared to survivors with musculoskeletal or gastrointestinal (MSK/GI) sequelae. Logistic regression performed in R Studio. PES= Post-Ebola syndrome, EVD= Ebola virus disease.

| | FC | log2(FC) | p.adjusted | -log10(p) |
|---|---|---|---|---|
| Acrylate | 4.0574 | 2.0206 | 1.83E-05 | 4.7381 |
| Acetate | 3.6465 | 1.8665 | 1.83E-05 | 4.7381 |
| 3-Hydroxy-3-methylglutarate | 3.6228 | 1.8571 | 1.83E-05 | 4.7381 |
| Glucose | 2.9963 | 1.5832 | 1.83E-05 | 4.7381 |
| Malate | 0.45773 | -1.1274 | 9.78E-05 | 4.0099 |
| 2-Oxo-4-hydroxy-4-carboxy-5-ureidoimidazoline | 4.6347 | 2.2125 | 0.00010336 | 3.9857 |
| Succinate | 0.45074 | -1.1496 | 0.00010336 | 3.9857 |
| Methylmalonate | 0.45074 | -1.1496 | 0.00010336 | 3.9857 |
| Threonate-1,4-lactone | 0.45074 | -1.1496 | 0.00010336 | 3.9857 |
| Propanoate | 0.48027 | -1.0581 | 0.00043485 | 3.3617 |
| Oxalosuccinate | 7.7896 | 2.9615 | 0.0016024 | 2.7952 |
| Palmitoylcarnitine | 0.38619 | -1.3726 | 0.0022608 | 2.6457 |
| Ascorbate | 0.16542 | -2.5958 | 0.0036576 | 2.4368 |
| N-Acetylornithine | 0.017035 | -5.8753 | 0.0068859 | 2.162 |
| Deoxycytidine | 0.14416 | -2.7942 | 0.0068859 | 2.162 |
| Hypoxanthine | 0.26767 | -1.9015 | 0.0076167 | 2.1182 |
| 2-Hydroxyhippurate | 0.38912 | -1.3617 | 0.0080669 | 2.0933 |
| Dopaquinone | 0.38912 | -1.3617 | 0.0080669 | 2.0933 |
| Leucodopachrome | 0.38912 | -1.3617 | 0.0080669 | 2.0933 |
| alpha-Hydroxyhippurate | 0.38912 | -1.3617 | 0.0080669 | 2.0933 |
| 4- Carboxyphenylglycine | 0.38912 | -1.3617 | 0.0080669 | 2.0933 |
| Dihydrouracil | 0.39851 | -1.3273 | 0.0080669 | 2.0933 |
| gamma-Glutamyl-2-aminobutyrate | 0.035676 | -4.8089 | 0.0084969 | 2.0707 |
| Cysteinylglycine | 0.23083 | -2.1151 | 0.0084969 | 2.0707 |
| 2-Oxoglutarate | 0.21137 | -2.2421 | 0.010247 | 1.9894 |
| Inosine-5'-monophosphate (IMP) | 0.4141 | -1.2719 | 0.014766 | 1.8307 |
| 6-Phospho-gluconate | 0.21403 | -2.2241 | 0.015814 | 1.801 |
| Carnosine | 0.25697 | -1.9603 | 0.024622 | 1.6087 |
| Diethanolamine | 2.7817 | 1.476 | 0.030025 | 1.5225 |
| Methyluridine | 0.38369 | -1.382 | 0.030025 | 1.5225 |
| Aspartate | 2.6075 | 1.3827 | 0.035226 | 1.4531 |
| 4-Fumarylacetoacetate | 0.49123 | -1.0255 | 0.039728 | 1.4009 |
| 4-Maleylacetoacetate | 0.49123 | -1.0255 | 0.039728 | 1.4009 |
| Uridine monophosphate (UMP) | 0.45329 | -1.1415 | 0.042379 | 1.3728 |

Supplementary table 3: Fold change (FC) and adjusted p-values (false discovery rate (FDR) from Benjamini-Hochberg (BH) method) for 34 significantly differential metabolites between EVD survivors with PES and asymptomatic survivors.

| Variable | by Variable | Spearman ρ | Prob>\|ρ\| | BH.corr.pvalue |
|---|---|---|---|---|
| Succinate | Glucose | -0.5960 | 0.0001 | 0.00065941 |
| Methylmalonate | Glucose | -0.5960 | 0.0001 | 0.00065941 |
| Methylmalonate | Succinate | 1 | 0.0001 | 0.00065941 |
| Succinate | Malate | 0.4407 | 0.0063 | 0.02370508 |
| Methylmalonate | Malate | 0.4407 | 0.0063 | 0.02370508 |
| Glucose | Malate | -0.4066 | 0.0125 | 0.04141791 |
| Threonate-1,4-lactone | Glucose | -0.5960 | 0.0001 | 0.00065941 |
| Threonate-1,4-lactone | Succinate | 1 | 0.0001 | 0.00065941 |
| Threonate-1,4-lactone | Methylmalonate | 1 | 0.0001 | 0.00065941 |
| Acrylate | Glucose | 0.9381 | 0.0001 | 0.00065941 |
| Deoxycytidine | Malate | 0.5861 | 0.0001 | 0.00065941 |
| N-Acetylornithine | Glucose | -0.6449 | 0.0001 | 0.00065941 |
| Inosine-5'-monophosphate (IMP) | Glucose | -0.6361 | 0.0001 | 0.00065941 |
| Propanoate | Succinate | 0.826 | 0.0001 | 0.00065941 |
| Propanoate | Methylmalonate | 0.826 | 0.0001 | 0.00065941 |
| 2-Oxo-4-hydroxy-4-carboxy-5-ureidoimidazoline | Glucose | 0.7892 | 0.0001 | 0.00065941 |
| 3-Hydroxy-3-methylglutarate | Glucose | 0.8888 | 0.0001 | 0.00065941 |
| Acetate | Glucose | 0.9263 | 0.0001 | 0.00065941 |
| Acetate | Succinate | -0.5846 | 0.0001 | 0.00065941 |
| Acetate | Methylmalonate | -0.5846 | 0.0001 | 0.00065941 |
| 6-Phospho-gluconate | Succinate | 0.6264 | 0.0001 | 0.00065941 |
| 6-Phospho-gluconate | Methylmalonate | 0.6264 | 0.0001 | 0.00065941 |
| Acrylate | Succinate | -0.5799 | 0.0002 | 0.00122202 |
| Acrylate | Methylmalonate | -0.5799 | 0.0002 | 0.00122202 |
| 2-Oxo-4-hydroxy-4-carboxy-5-ureidoimidazoline | Succinate | -0.5778 | 0.0002 | 0.00122202 |
| 2-Oxo-4-hydroxy-4-carboxy-5-ureidoimidazoline | Methylmalonate | -0.5778 | 0.0002 | 0.00122202 |
| N-Acetylornithine | Succinate | 0.5614 | 0.0003 | 0.00175263 |
| N-Acetylornithine | Methylmalonate | 0.5614 | 0.0003 | 0.00175263 |
| 3-Hydroxy-3-methylglutarate | Succinate | -0.5372 | 0.0006 | 0.00327541 |
| 3-Hydroxy-3-methylglutarate | Methylmalonate | -0.5372 | 0.0006 | 0.00327541 |
| 2-Hydroxyhippurate | Succinate | 0.5332 | 0.0007 | 0.00335396 |
| 2-Hydroxyhippurate | Methylmalonate | 0.5332 | 0.0007 | 0.00335396 |
| Dopaquinone | Succinate | 0.5332 | 0.0007 | 0.00335396 |

| | | | | |
|---|---|---|---|---|
| Dopaquinone | Methylmalonate | 0.5332 | 0.0007 | 0.00335396 |
| Leucodopachrome | Succinate | 0.5332 | 0.0007 | 0.00335396 |
| Leucodopachrome | Methylmalonate | 0.5332 | 0.0007 | 0.00335396 |
| alpha-Hydroxyhippurate | Succinate | 0.5332 | 0.0007 | 0.00335396 |
| alpha-Hydroxyhippurate | Methylmalonate | 0.5332 | 0.0007 | 0.00335396 |
| 4- Carboxyphenylglycine | Succinate | 0.5332 | 0.0007 | 0.00335396 |
| 4- Carboxyphenylglycine | Methylmalonate | 0.5332 | 0.0007 | 0.00335396 |
| 6-Phospho-gluconate | Glucose | -0.4844 | 0.0024 | 0.01031226 |
| 2-Oxo-4-hydroxy-4-carboxy-5-ureidoimidazoline | Malate | -0.4704 | 0.0033 | 0.01382264 |
| Deoxycytidine | Glucose | -0.4535 | 0.0048 | 0.01891598 |
| Uridine monophosphate (UMP) | Glucose | -0.4538 | 0.0048 | 0.01891598 |
| Threonate-1,4-lactone | Malate | 0.4407 | 0.0063 | 0.02370508 |
| 6-Phospho-gluconate | 2-Oxoglutarate | -0.4391 | 0.0066 | 0.02455642 |
| Cysteinylglycine | Malate | 0.4365 | 0.0069 | 0.02538895 |
| 3-Hydroxy-3-methylglutarate | Malate | -0.4367 | 0.0069 | 0.02538895 |
| Hypoxanthine | Glucose | -0.4334 | 0.0074 | 0.02693115 |
| 3-Hydroxy-3-methylglutarate | Oxalosuccinate | 0.4294 | 0.008 | 0.02895652 |
| Acrylate | Malate | -0.4286 | 0.0081 | 0.02900323 |
| 2-Oxo-4-hydroxy-4-carboxy-5-ureidoimidazoline | Oxalosuccinate | 0.4229 | 0.0091 | 0.03156563 |
| Acetate | Malate | -0.4229 | 0.0091 | 0.03156563 |
| Acrylate | Oxalosuccinate | 0.408 | 0.0122 | 0.040626 |
| Acetate | Oxalosuccinate | 0.3988 | 0.0145 | 0.04710732 |
| Glucose | Diethanolamine | 0.674 | 0.0001 | 0.00065941 |
| Succinate | gamma-Glutamyl-2-aminobutyrate | 0.5863 | 0.0001 | 0.00065941 |
| Methylmalonate | gamma-Glutamyl-2-aminobutyrate | 0.5863 | 0.0001 | 0.00065941 |
| 2-Oxoglutarate | Ascorbate | 0.811 | 0.0001 | 0.00065941 |
| 2-Oxoglutarate | 4-Fumarylacetoacetate | 0.7044 | 0.0001 | 0.00065941 |
| 2-Oxoglutarate | 4-Maleylacetoacetate | 0.7044 | 0.0001 | 0.00065941 |
| Malate | Ascorbate | 0.5538 | 0.0004 | 0.00225763 |
| Glucose | gamma-Glutamyl-2-aminobutyrate | -0.5555 | 0.0004 | 0.00225763 |
| 2-Oxoglutarate | Carnosine | -0.5344 | 0.0007 | 0.00335396 |
| Succinate | Carnosine | 0.4919 | 0.002 | 0.00888 |
| Methylmalonate | Carnosine | 0.4919 | 0.002 | 0.00888 |
| Succinate | Diethanolamine | -0.4689 | 0.0034 | 0.01389202 |

| | | | | |
|---|---|---|---|---|
| Methylmalonate | Diethanolamine | -0.4689 | 0.0034 | 0.01389202 |
| Glucose | Carnosine | -0.4244 | 0.0089 | 0.0313619 |

Supplementary Table 4: Spearman ρ values for metabolites related to TCA cycle or glucose. Metabolites considered related to the TCA cycle and that had significant interactions included succinate, malate, methylmalonate, oxalosuccinate, 2-oxoglutarate. Only correlations with a BH corrected p value of <.05 were included.

| Component 1 | | |
|---|---|---|
| **Metabolite** | **VIP Score** | **Significance - PES** |
| Acetate | 2.6838 | Upregulated |
| Acrylate | 2.6663 | Upregulated |
| Glucose | 2.6593 | Upregulated |
| 3-Hydroxy-3-methylglutarate | 2.6552 | Upregulated |
| Malate | 2.5089 | Downregulated |
| 2-Oxo-4-hydroxy-4-carboxy-5-ureidoimidazoline | 2.4658 | Upregulated |
| Succinate | 2.4551 | Downregulated |
| Methylmalonate | 2.4551 | Downregulated |
| Threonate-1,4-lactone | 2.4551 | Downregulated |
| Propanoate | 2.3163 | Downregulated |
| LysoPA(16:0/0:0) | 2.2934 | Not significant |
| Lyxose | 2.1664 | Not significant |
| Oxalosuccinate | 2.1586 | Upregulated |

| | | |
|---|---|---|
| Tyramine | 2.1061 | Not significant |
| Phenylethanolamine | 2.1061 | Not significant |
| **Component 2** | | |
| **Metabolite** | **VIP Score** | **Significance - PES** |
| 4-Hydroxy-enol-phenylpyruvate | 2.3796 | Not significant |
| 4-Hydroxyphenylpyruvate | 2.3796 | Not significant |
| Caffeate | 2.3796 | Not significant |
| Acetate | 2.3736 | Upregulated |
| Acrylate | 2.3626 | Upregulated |
| Glucose | 2.3488 | Upregulated |
| 3-Hydroxy-3-methylglutarate | 2.3391 | Upregulated |
| Malate | 2.2013 | Downregulated |
| Succinate | 2.1881 | Downregulated |
| Methylmalonate | 2.1881 | Downregulated |
| Threonate-1,4-lactone | 2.1881 | Downregulated |
| 2-Oxo-4-hydroxy-4-carboxy-5-ureidoimidazoline | 2.1641 | Upregulated |
| Palmitoylcarnitine | 2.125 | Downregulated |
| LysoPA(16:0/0:0) | 2.1045 | Not significant |
| Propanoate | 2.0713 | Downregulated |

Supplementary Table 5: Top 15 variables of importance (VIP) for PLS-DA model. Metabolite considered significantly upregulated or downregulated if FC > 2.0 and FDR <.05 when comparing PES and asymptomatic survivor groups.

**Supplementary Figures**

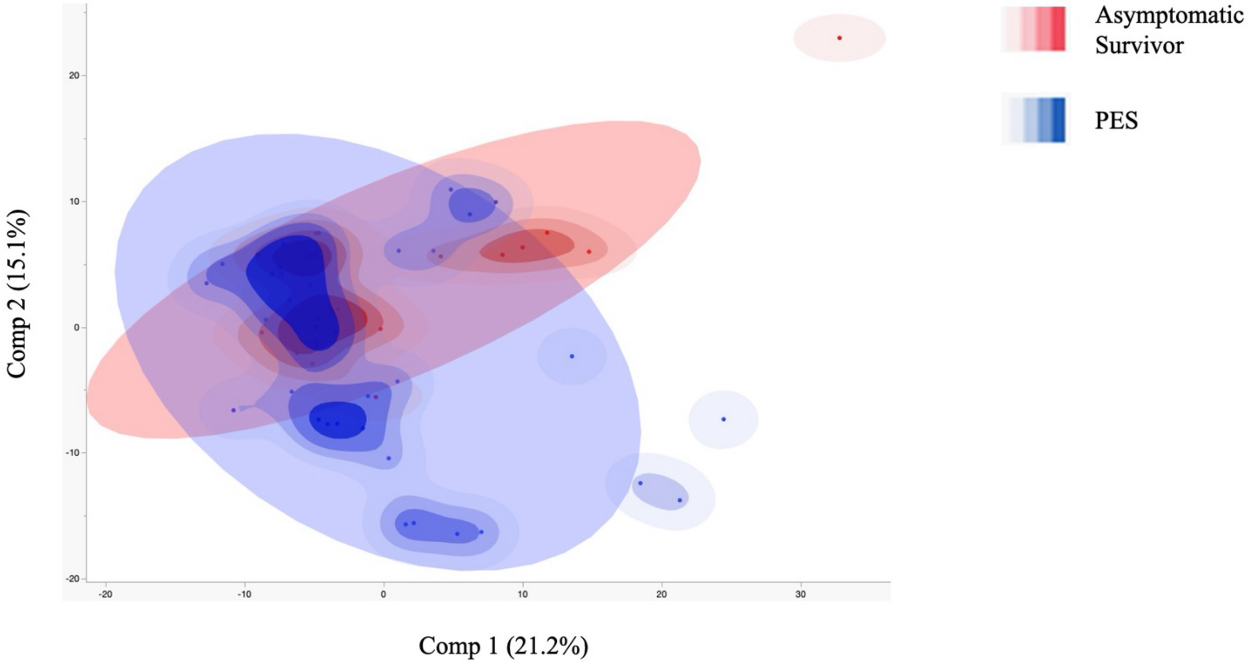

Supplementary Figure 1: Principal Components Analysis (PCA) Demonstrates Separation of Survivors with PES and Asymptomatic Survivors

EVD survivors with PES separate from asymptomatic survivors based on metabolic profile. Data analyzed in MetaboAnalyst and visualized in JMP.

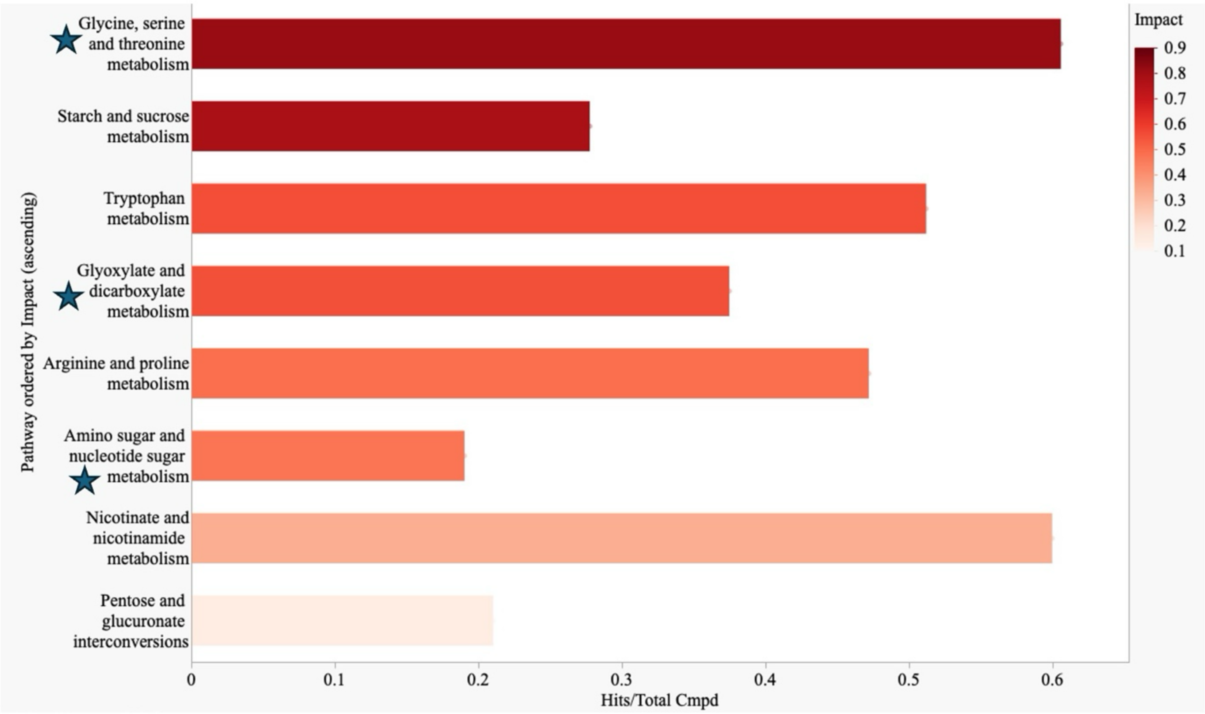

Supplementary Figure 2: Several Metabolic Pathways are Significantly Upregulated or Tied in EVD Survivors with PES Compared to Asymptomatic Survivors

Pathways with an equal number of upregulated and downregulated ("tie" pathways) metabolites are denoted by a star. Pathways are ordered by impact score. Impact score required to be >0.2 and FDR <.05 for inclusion. Pathway analysis performed in MetaboAnalyst and data visualized using JMP.

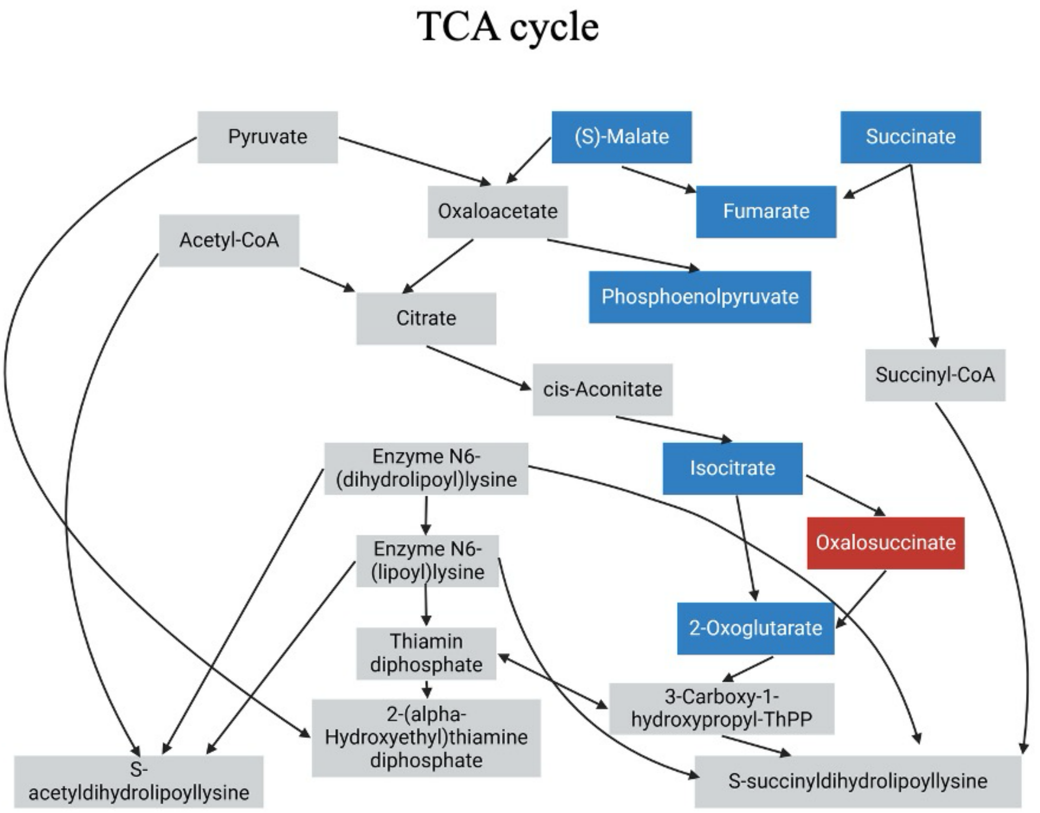

Supplementary Figure 3: EVD Survivors with PES Demonstrates Primarily Downregulated Metabolism of the TCA Cycle Compared to Asymptomatic Survivors

Significantly downregulated metabolites are highlighted in blue. Significantly upregulated metabolites are highlighted in red. Metabolites that were neither significantly upregulated or downregulated are in grey. Pathway analysis performed in MetaboAnalyst. Created in BioRender. Sanford, A. (2026) https://BioRender.com/v1jj72h

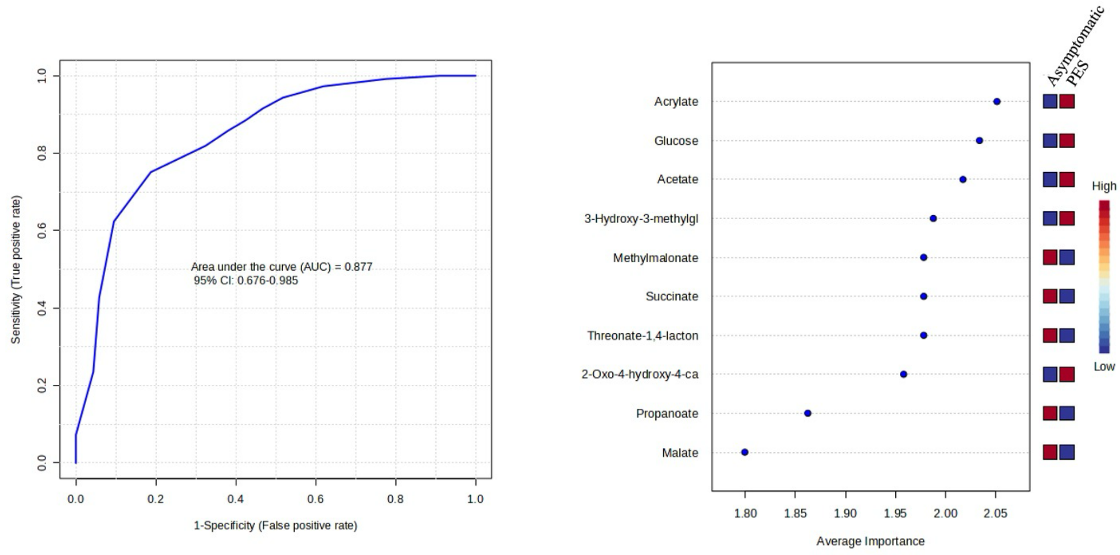

A

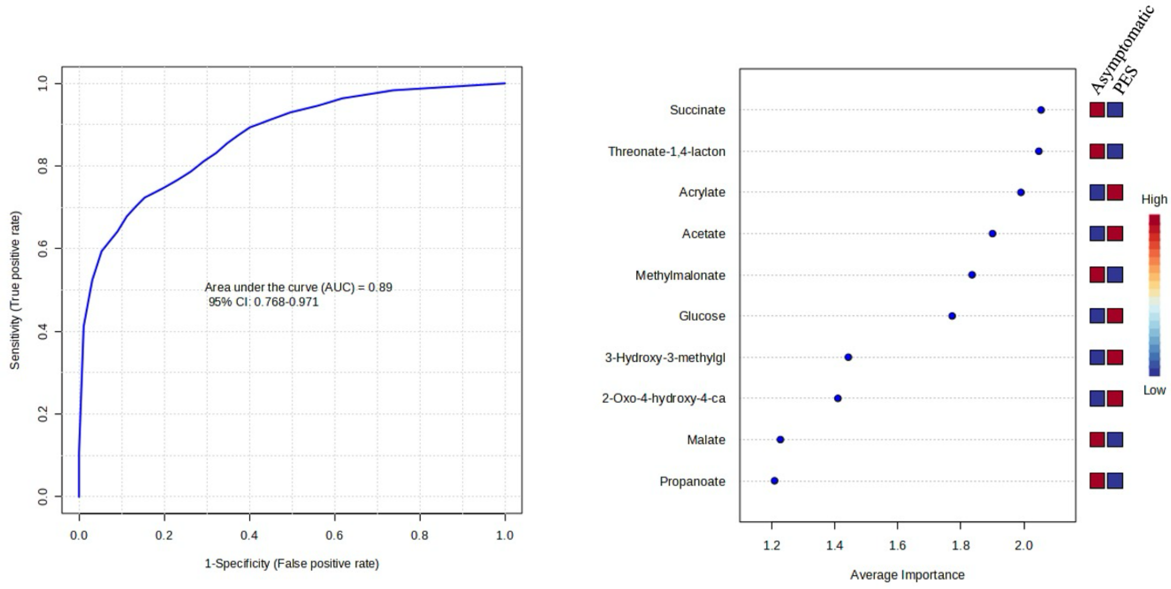

B

Supplementary Figure 4: Discovery Models of Biomarkers of PES

Biomarker analysis was performed to identify group membership (PES vs asymptomatic survivor) in MetaboAnalyst. Discovery (n= 39) and validation (n= 18) cohorts were used (discovery cohorts shown here, validation cohorts in Figures 5A-B). Potential biomarkers were selected from PLS-DA and Random Forest (RF) modeling in the discovery cohort. The top 10 most important features for predicting PES were the same in RF and PLS-DA modeling. If the metabolite was higher in the PES or asymptomatic group, the box to the right of the figure was marked red, and if that metabolite was lower, the box was marked blue. Figure 4A:

ROC from discovery model from PLS-DA and 10 most important features, ranked by importance. Figure 4B: ROC from discovery model from RF and 10 most important features, ranked by importance.
